# Machine Learning-Based Identification of Patients with Elevated Central Venous Pressure Using Features Extracted from Photoplethysmography Waveforms

**DOI:** 10.64898/2025.12.30.25343231

**Authors:** Ravi Pal, Akos Rudas, Jeffrey N. Chiang, Anna Barney, Maxime Cannesson

## Abstract

Central venous pressure (CVP), a key component of hemodynamic monitoring, is widely used to guide fluid resuscitation in critically ill patients. It is typically measured using central venous line catheterization, which is the gold standard, but this method is invasive, time-consuming, and associated with complications. This study aims to investigate whether machine learning (ML)-based analysis of features extracted from a non-invasive, standard-of-care waveform—the photoplethysmography (PPG) signal—can identify patients with elevated CVP. We trained Light Gradient-Boosting Machine (LightGBM) model using a large perioperative dataset (MLORD), containing 17,327 surgical patients from 2019 to 2022 at UCLA. For this study, we selected 1665 patients with both PPG and CVP waveforms available. A total of 843 PPG features per cardiac cycle (CC) were extracted from the PPG waveforms using a signal processing-based feature extraction tool, along with the simultaneous maximum value calculated from the corresponding CCs in the CVP waveform. Additionally, for each patient, the average and standard deviation of each PPG feature, as well as the mean of the maximum CVP values, were calculated across all cardiac cycles, resulting in 843 averaged PPG features, 843 PPG feature standard deviations, and one averaged maximum CVP value per patient. The average maximum CVP value was used as the ground truth to classify patients as either normal (5 ≤ CVP ≤ 15 mmHg) or elevated (CVP > 15 mmHg). Of the 1,665 patients, 1,182 were normal and 483 were elevated. The dataset was split into 90% for training (1,063 normal and 435 elevated) and 10% for testing (119 normal and 48 elevated). From the 1686 PPG features (843 averaged and 843 standard deviation), 246 were selected for model development using the Recursive Feature Elimination with Cross-Validation (RFECV) approach. To further enhance performance, hyperparameters were tuned through 5-fold cross-validation on the training set. Finally, the best-performing configuration was retrained on the full training data, and its performance was evaluated on the held-out test set. To provide a robust estimate and confidence interval, a bootstrapping procedure with 100 iterations was performed on the test set. The LightGBM classifier achieved a mean area under the receiver operating characteristic curve (AUC) of 0.79 (95% CI: 0.71–0.84) and mean accuracy of 0.71 (95% CI: 0.65-0.77), demonstrating good discriminatory power in distinguishing between patients with normal and elevated CVP. This study highlights the ability of PPG-derived features to discriminate between patients with normal and elevated CVP using ML. These early findings lay the groundwork for future research aimed at developing non-invasive approaches to CVP assessment.

## Introduction

Human biosignals such as photoplethysmography (PPG) and central venous pressure (CVP) waveforms carry important physiological information crucial for diagnosing and monitoring patients’ health [1,2,3]. In this study, we investigate whether features derived from PPG waveforms can discriminate between patients with normal and elevated CVP using machine learning (ML). The scientific basis of our hypothesis is that the venous pulse can be identified in the PPG waveform, and its presence, especially at higher CVP, affects the waveform’s shape [4,5,6,7,8,9]. Therefore, the PPG waveform analyzed using advanced signal processing and ML can provide a non-invasive way of CVP assessment.

CVP measurement is a widely used method in operating rooms and intensive care units for estimating the body’s need for fluids in patients with hemodynamic instability [10,11,12]. The clinical range of CVP is generally 5 to 15 mmHg [13]. It is considered a reliable indicator of right ventricular preload, used to assess cardiac preload and volume status, and supports the diagnosis of right-sided heart failure [14,15]. It is typically measured by inserting a catheter into the upper vena cava via the internal jugular vein or subclavian [12]. However, this approach is invasive and can be time-consuming [16]. In addition, it carries potential risks such as vascular injury and requires skilled personnel to perform the procedure safely [16,17].

In contrast, PPG is a non-invasive, standard-of-care signal used in clinical settings to measure blood oxygen saturation through pulse oximetry [1,18]. The abundance of physiological information it provides, combined with its ease of use, has led to its application beyond this primary purpose. PPG has been widely applied to estimate physiological parameters and detect various conditions, including blood pressure, atrial fibrillation, diabetes, vascular aging, sleep disorders (such as apnea and hypopnea), emotional states, and cardiac output [1,19]. Moreover, with recent advancements in signal processing and the rise of ML, PPG signal analysis has become increasingly integrated into wearable health devices such as smartwatches and fitness trackers [20,21, 22]. These technologies—signal processing and ML—have opened new avenues for extracting clinically relevant information from PPG, enabling everyday health monitoring—for example, heart rate tracking [21,23].

In the context of CVP analysis using PPG waveform, studies have shown that variations in the PPG waveform can reflect peripheral venous pulsations influenced by CVP [4]. Additionally, reflectance-mode contact PPG applied to the anterior neck has shown promise for capturing the jugular venous pressure waveform, which can be used to estimate CVP [24]. However, despite PPG’s widespread availability and noninvasive nature, its application for CVP assessment remains relatively underexplored.

Therefore, this paper aims to explore the utilization of features extracted from PPG waveforms for CVP analysis using ML. We applied our recently developed feature extraction tool [25,26] to derive features from PPG waveforms. Using these features, we trained a Light Gradient-Boosting Machine (LightGBM) ML classifier [27] to differentiate between patients with normal and elevated CVP. Additionally, we employed SHapley Additive exPlanations (SHAP) [28] to explain the output of the ML model. SHAP analysis provides information about how much each input feature (in our study, a PPG feature) contributes to the model output [29]. While this study does not directly estimate the CVP waveform from the non-invasive PPG signal, it lays the groundwork for future research into non-invasive CVP assessment techniques.

The key contributions of the study are:

1. Examination using a LightGBM classifier of the potential for PPG-derived features—including time-domain, frequency-domain, and statistical features—to identify patients with elevated CVP using ML.
2. Application of an explainable AI approach, SHAP, to the model output to evaluate the contribution of individual PPG features to the model’s identification of patients with elevated CVP.
3. Rigorous evaluation of the model’s performance on a test dataset to quantify robustness of classification performance.

## Methods

Previous studies have shown that the morphology of the PPG waveform may be influenced by venous pressure [4,7,24], with the diastolic phase of the PPG signal particularly amplified due to the venous component [4,7]. Therefore, under normal versus high CVP conditions, the PPG waveform may exhibit distinct morphological differences, especially in the diastolic phase. For example, Fig. 1(i-a) and Fig. 1(ii-a) illustrate differences in the diastolic phase of the PPG cardiac cycles corresponding to the high and normal CVP cycles shown in Fig. 1(i-b) and Fig. 1(ii-b), respectively. This difference is not always apparent in the raw PPG waveform, but advanced signal-processing–based feature extraction tools may extract clinically useful features, which, when used with ML models, can play a crucial role in identifying patients with elevated CVP.

**Fig. 1.**
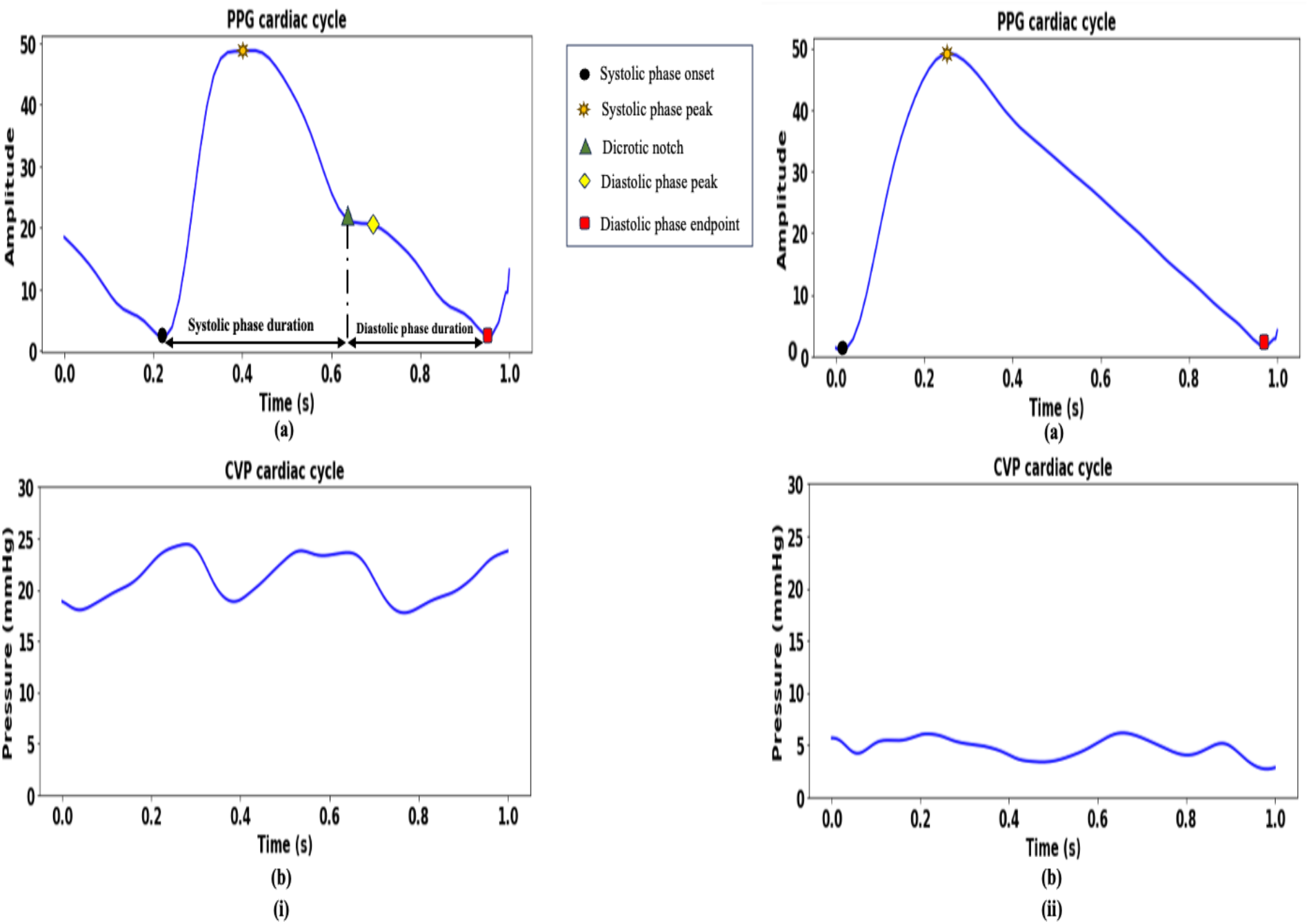
(i) Photoplethysmography (PPG) cardiac cycle with all five key landmarks (a) and the corresponding central venous pressure (CVP) cardiac cycle with high CVP (b); and (ii) PPG cardiac cycle with only three key landmarks visible (a) and the corresponding CVP cardiac cycle with normal CVP (b).

### Dataset

We utilized a large perioperative dataset (MLORD) containing physiological waveforms from 17,327 patients who underwent surgeries between 2019 and 2022 at the David Geffen School of Medicine, University of California Los Angeles (IRB# 19–000354; NIH research funding: R01HL14469) [30], to evaluate the performance of the ML model. The MLORD dataset includes both clinical and physiological waveform data. Clinical data were extracted from the electronic health record (EHR) platforms Epic (Verona, WI, USA) and Surgical Information Systems (Alpharetta, GA, USA). Waveform data were captured in the operating rooms using the Bernoulli data acquisition system (Cardiopulmonary, New Haven, CT, USA). In total, the waveform data contains more than 72,264 hours of recordings—approximately 7.6 TB—encompassing digital physiological waveforms such as electrocardiograms (ECG), arterial blood pressure (ABP), PPG, and CVP [30]. For this study, we focused on a subset of 1,665 patients for whom both PPG and CVP recordings were available, each sampled at 256 Hz.

### Waveform Feature Extraction

We used our recently developed signal processing–based feature extraction tool for extracting features from PPG waveforms [26]. The tool operates in two steps: first, it identifies all key landmarks within a cardiac cycle (systolic phase onset, systolic peak, dicrotic notch, diastolic peak, and diastolic endpoint), these key landmarks are shown in Figure 1(i-a); second, using these landmarks, it extracts more than 840 features per cycle. We selected this tool for PPG waveforms featurization for two main reasons: (1) its high accuracy in detecting key landmarks even when they are less visible in the input signal and extracting a large number of features, and: (2) to evaluate the clinical usability of the features the tool extracts. In this study, the tool extracted a total of 843 PPG features per cardiac cycle, including time-domain, frequency-domain, and statistical features. Separately, from the CVP waveform, the maximum value corresponding to each cardiac cycle was determined. These cardiac cycle–level features were then summarized at the patient level by calculating the mean and standard deviation of each PPG feature across all cycles, resulting in 843 mean PPG features and 843 PPG feature standard deviations per patient. The maximum CVP values were also averaged across all cardiac cycles to obtain a single mean maximum CVP value for each patient. For a detailed explanation of the feature extraction tool’s working process and a complete description of the PPG features, we refer readers to our previous articles [25,26].

### Model development

To prepare the dataset, it was divided into training and test subsets. In clinical practice, a CVP range of 5–15 mmHg is generally considered normal [13]. Accordingly, in this study, patients with a mean maximum CVP within this range were classified as having normal CVP and those with values above 15 mmHg were classified as having elevated CVP. Among the 1,665 patients analyzed, 1,182 were in the normal group and 483 in the elevated group. 90% (normal CVP: 1063 and elevated CVP: 435) of the data from both groups was used for model training and 10% (normal CVP: 119 and elevated CVP: 48) was reserved for independent testing. Feature selection was performed using Recursive Feature Elimination with Cross-Validation (RFECV) [31], which identified 246 informative features out of the 1686 PPG features (843 averaged and 843 standard deviation).

We developed a LightGBM classifier, selected for its performance and speed [27], and trained it using these informative PPG features to identify patients with elevated CVP. Hyperparameter tuning was performed using a grid search with 5-fold cross-validation on the training set to determine the optimal configuration. Once the best-performing settings were identified, the model was trained on the complete training set and evaluated on the held-out test set. Model performance was assessed using the area under the receiver operating characteristic curve (AUC) and accuracy. To ensure robustness, a 100-iteration bootstrapping procedure was applied to the test set to estimate performance metrics and their corresponding 95% confidence intervals [32].

### Interpretability analyses

We applied the SHapley Additive exPlanations (SHAP) method [28] to quantify the contribution of each PPG feature to the model’s predictions. SHAP is a widely used feature-based interpretability technique that can be integrated into ML models to improve transparency, trust, and understanding of their predictions [33]. The core idea of SHAP is to assign a Shapley value to each feature, reflecting its contribution to the model’s output for a given data point [29]. We conducted SHAP analysis using the Python SHAP package [34]. For clarity, a block diagram of the complete workflow used in this study is shown in Figure 2.

**Fig. 2.**
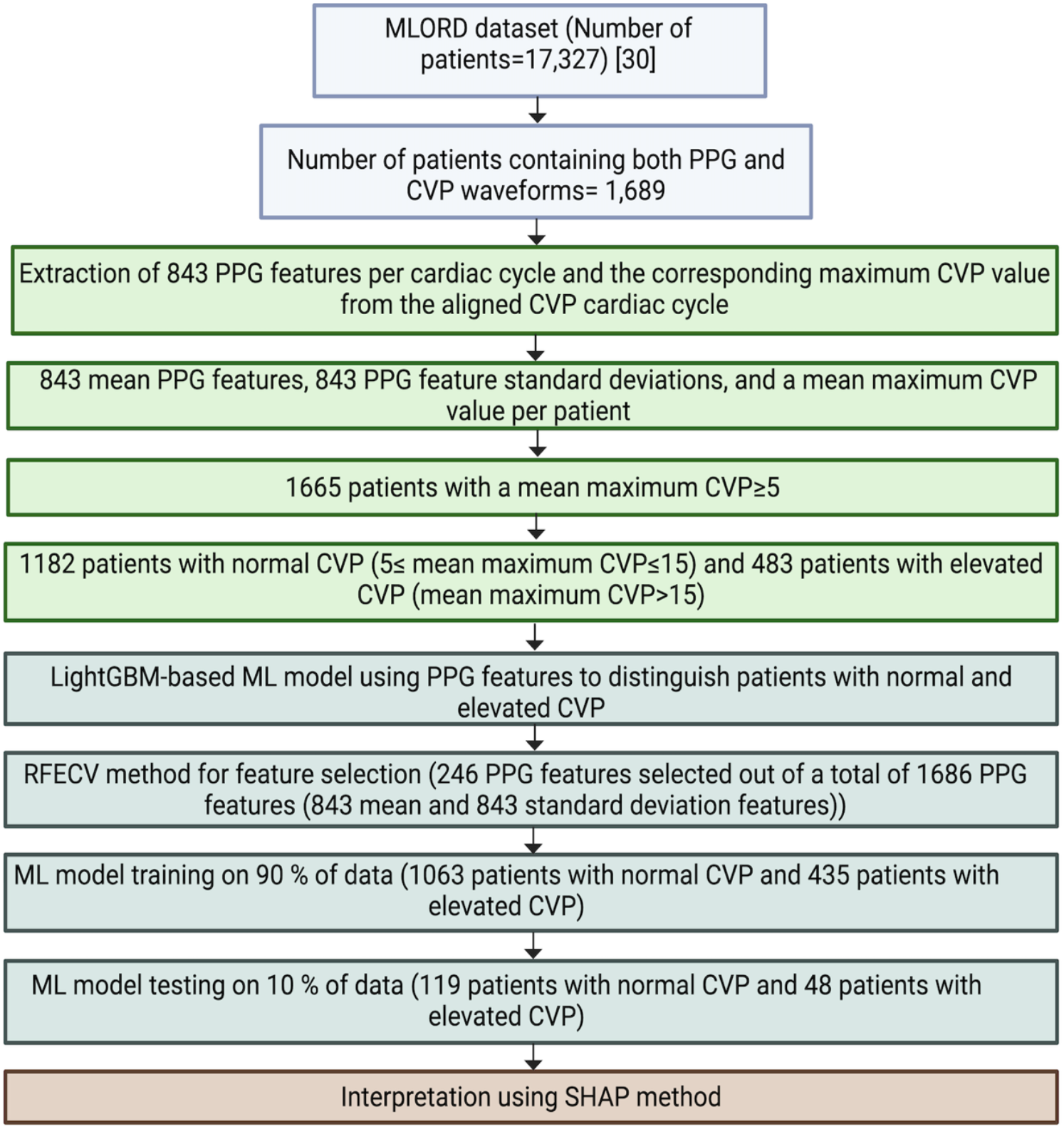
Block diagram of the workflow used in this study using the MLORD dataset.

## Results

### Model Performance

Model performance was evaluated using both the receiver operating characteristic (ROC) curve and accuracy [35]. On the test set, the LightGBM classifier demonstrated acceptable discriminative ability in distinguishing patients with elevated CVP from those with normal CVP, achieving a mean area under the ROC curve of 0.79 (95% CI: 0.71–0.84) and a mean accuracy of 0.71 (95% CI: 0.65, 0.77). The mean ROC curve with the corresponding 95% confidence interval is presented in Figure 3.

**Fig. 3.**
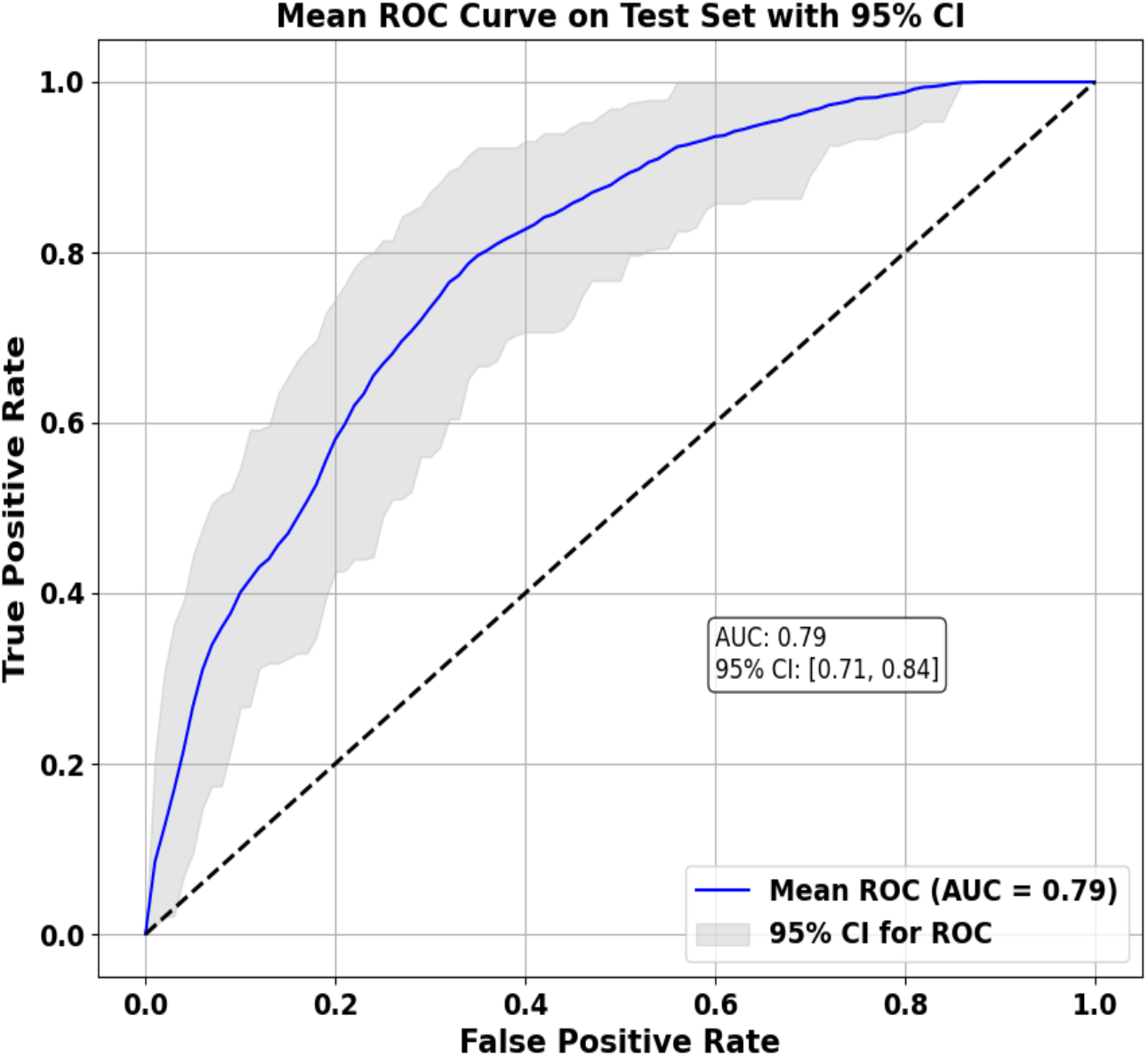
The receiver operating characteristic curve of the LightGBM model.

### SHAP method to explain the LightGBM classifications

The SHAP method was used to determine the contribution of each PPG feature to the prediction results of the LightGBM model. Figure 4a displays the SHAP feature importance bar plot, listing the top 20 most important PPG features in descending order, with features higher on the y-axis indicating a greater influence on model predictions. Figure 4b presents the SHAP summary plot, which depicts the distribution of SHAP values for these features across all samples, highlighting both the magnitude and direction of their effects on predictions. A positive SHAP value indicates an increased risk of elevated CVP while negative value suggests a decreased risk. In this plot, each point represents an individual sample, and the colour represents the associated feature value, with high values shown in red and low values in blue, thereby providing insight into how variation in feature values affects the model’s output. A complete description of all top 20 PPG features is provided in Table 1.

**Table 1.** Detailed descriptions of the top 20 most important PPG features that contributed to the ML model prediction (Fig. 4).

|  | Feature | Description |
| --- | --- | --- |
| 1 | PPG_pkfrq_std | The standard deviation of the frequency corresponding to the highest peak in the FFT of the PPG cardiac cycle |
| 2 | PPG_W_SRP33_std | The standard deviation of the PPG cardiac cycle systolic rise phase width at 33% of the cardiac cycle |
| 3 | PPG_AM_RT_SD_Cpeak_wrtDpeak_PPG_AM_SD_Dpeak_wrtzero_std | The standard deviation of the ratio of the PPG cardiac cycle second derivative Cpeak amplitude with respect to Dpeak to the PPG cardiac cycle second derivative Dpeak amplitude with respect to zero |
| 4 | PPG_NAM_RT_DIAPE_wrtZero_PPG_DIAN_wrtDIAPE_std | The standard deviation of the ratio of the normalized PPG cardiac cycle diastolic phase end point amplitude with respect to zero to the normalized PPG cardiac cycle diastolic notch amplitude with respect to diastolic phase end point |
| 5 | PPG_AM_RT_SD_Cpeak_wrtzero_PPG_AM_SD_Bpeak_wrtzero_std | The standard deviation of the ratio of the PPG cardiac cycle second derivative Cpeak amplitude with respect to zero to the PPG cardiac cycle second derivative Bpeak amplitude with respect to zero |
| 6 | PPG_AM_RT_SD_Cpeak_wrtEpeak_PPG_AM_SD_Dpeak_wrtzero_mean | The mean of the ratio of the PPG cardiac cycle second derivative Cpeak amplitude with respect to Epeak to the PPG cardiac cycle second derivative Dpeak amplitude with respect to zero |
| 7 | PPG_D_SD_Apeak_wrtDpeak_mean | The mean of the duration of the PPG cardiac cycle second derivative Apeak duration with respect to Dpeak |
| 8 | PPG_AM_RT_FD_Vpeak_wrtWpeak_PPG_AM_FD_Wpeak_wrtzero_std | The standard deviation of the ratio of the PPG cardiac cycle first derivative Vpeak amplitude with respect to Wpeak to the PPG cardiac cycle first derivative Wpeak amplitude with respect to zero |
| 9 | PPG_AM_RT_FD_Upeak_wrtWpeak_PPG_AM_FD_Vpeak_wrtzero_std | The standard deviation of the ratio of the PPG cardiac cycle first derivative Upeak amplitude with respect to Wpeak to the PPG cardiac cycle first derivative Vpeak amplitude with respect to zero |
| 10 | PPG_AM_SD_Bpeak_wrtEpeak_std | The standard deviation of the PPG cardiac cycle second derivative Bpeak amplitude with respect to Epeak |
| 11 | PPG_AM_RT_SYSP_wrtDIAN_PPG_SYSP_wrtZero_std | The standard deviation of the ratio of the PPG cardiac cycle systolic phase peak amplitude with respect to diastolic notch to the PPG cardiac cycle systolic phase onset amplitude with respect to zero |
| 12 | PPG_NAM_SYSP_wrtDIAN_std | The standard deviation of the normalized PPG cardiac cycle systolic phase peak amplitude with respect to diastolic notch |
| 13 | PPG_AM_RT_DIAPE_wrtSYSP_PPG_DIAP_wrtDIAPE_mean | The mean of the ratio of the PPG cardiac cycle diastolic phase end point amplitude with respect to systolic phase onset to the PPG cardiac cycle diastolic phase peak amplitude with respect to diastolic phase end point |
| 14 | PPG_AM_RT_SD_Cpeak_wrtEpeak_PPG_AM_SD_Cpeak_wrtDpeak_std | The standard deviation of the ratio of the PPG cardiac cycle second derivative Cpeak amplitude with respect to Epeak to the PPG cardiac cycle second derivative Cpeak amplitude with respect to Dpeak |
| 15 | PPG_D_Avg_SYSP_wrtDIAPE_mean | The mean of the average duration of the PPG cardiac cycle systolic phase onset with respect to diastolic phase end point |
| 16 | PPG_AM_RT_DIAP_wrtDIAN_PPG_DIAP_wrtDIAPE_mean | The mean of the ratio of the PPG cardiac cycle diastolic phase peak amplitude with respect to diastolic notch to the PPG diastolic phase peak with respect to diastolic phase end point |
| 17 | PPG_D_SD_Cpeak_wrtDpeak_mean | The mean of the duration of the PPG cardiac cycle second derivative Cpeak duration with respect to Dpeak |
| 18 | PPG_NAM_RT_DIAPE_wrtSYSP_PPG_DIAPE_wrtZero_std | The standard deviation of the ratio of the normalized PPG cardiac cycle diastolic phase end point amplitude with respect to systolic phase onset to the normalized PPG cardiac cycle diastolic phase end point amplitude with respect to zero |
| 19 | PPG_AM_RT_DIAPE_wrtSYSP_PPG_DIAP_wrtDIAN_std | The standard deviation of the ratio of the PPG cardiac cycle diastolic phase end point amplitude with respect to systolic phase onset to the PPG cardiac cycle diastolic phase peak amplitude with respect to diastolic notch |
| 20 | PPG_D_RT_SYSP_wrtDIAPE_PPG_SYSP_wrtSYSP_std | The standard deviation of the ratio of the PPG cardiac cycle systolic phase peak duration with respect to diastolic phase end point to the PPG cardiac cycle systolic phase onset duration with respect to systolic phase peak |

PPG: Photoplethysmography, FFT: Fast Fourier Transform. \_mean: mean across cardiac cycles for each subject; \_std: standard deviation across cardiac cycles for each subject.

**Fig. 4.**
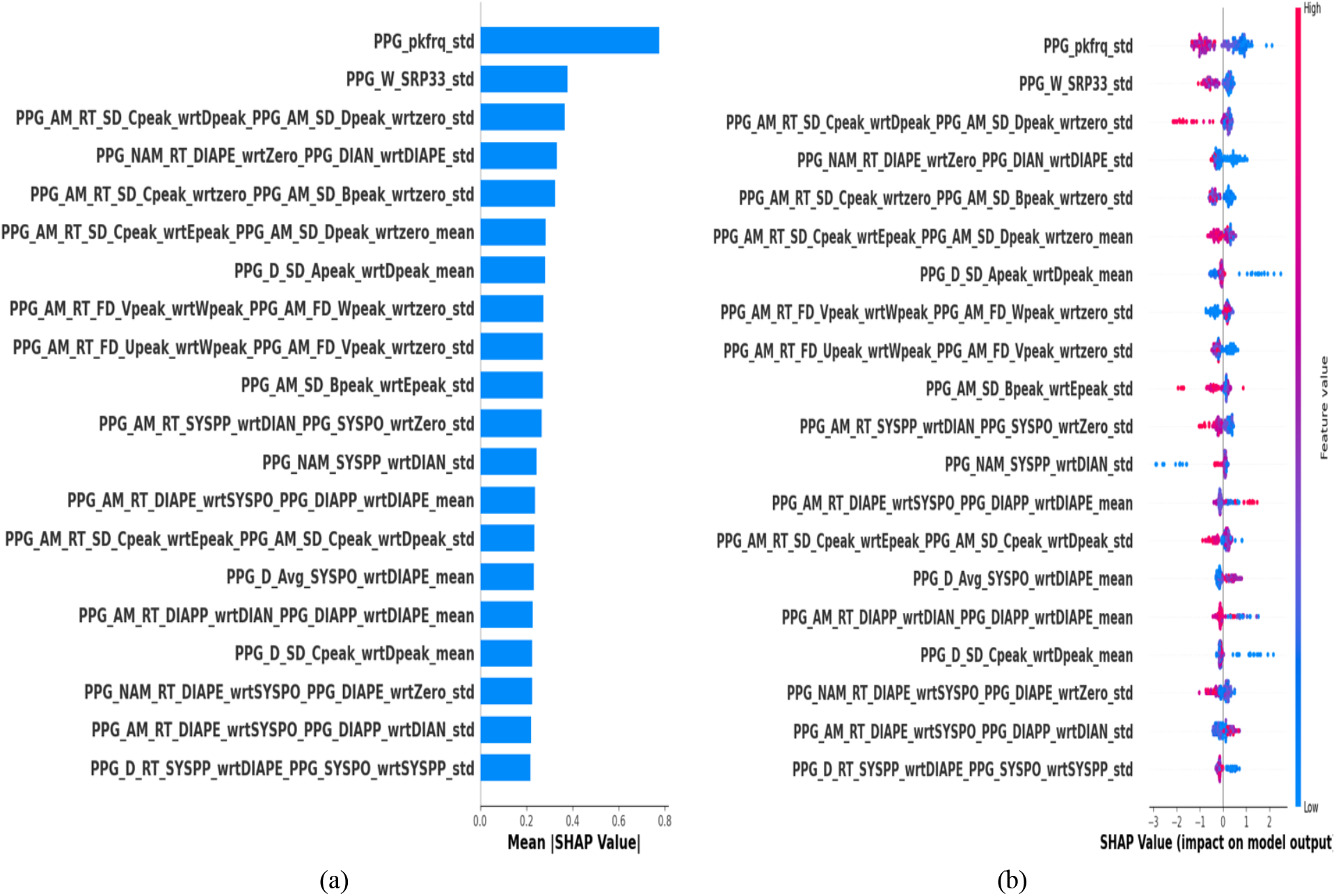
(a) SHAP feature importance bar plot of the top 20 PPG features in the LightGBM model; (b) SHAP summary plot of these PPG features. Features are ranked in descending order of importance in the model.

Notably, several PPG feature variability measures showed a significant influence on model prediction, as revealed by the SHAP analysis (Figure 4). These included the standard deviation of the dominant frequency of the PPG cardiac cycle, the standard deviation of the PPG cardiac cycle systolic rise phase width at 33% of the cardiac cycle, the standard deviation of the ratio of the PPG cardiac cycle second derivative Cpeak amplitude with respect to Dpeak to the PPG cardiac cycle second derivative Dpeak amplitude with respect to zero, the standard deviation of the ratio of the normalized PPG cardiac cycle diastolic phase end point amplitude with respect to zero to the normalized PPG cardiac cycle dicrotic notch amplitude with respect to diastolic phase end point, and the standard deviation of the ratio of the PPG cardiac cycle second derivative Cpeak amplitude with respect to zero to the PPG cardiac cycle second derivative Bpeak amplitude with respect to zero, showed significance influence on model prediction, highlighting, through the SHAP analysis, their importance in identifying elevated CVP.

## Discussion

Elevated CVP is often associated with organ dysfunction [36], a non-invasive approach for identifying patients with elevated CVP could facilitate the timely management of their hemodynamic instability. PPG has been suggested as a potential estimator of CVP [37]; however, research into PPG-based assessment of CVP remains limited. In this study, we examined for the first time the potential of ML–based analysis of PPG waveform features to identify patients with elevated CVP. This study utilized a high-performance ML model, LighGBM, for this task [27].

PPG waveforms are non-invasive and rich in physiological information. This has made them a focus of research across diverse clinical applications [19], ranging from vital sign estimation (such as pulse rate, blood pressure, and respiratory rate) to more complex diagnostic applications like sleep analysis [18]. However, Charlton et al. [23] highlighted that selecting an appropriate analytical technique is critical for PPG waveform analysis, as it can significantly affect the accuracy and precision of the extracted features. Therefore, in this study, we used our recently developed feature extraction tool to analyse PPG waveforms. As mentioned in Section 2, this tool accurately identifies key landmarks within each cardiac cycle and extracts a comprehensive set of features (over 840 per cardiac cycle), encompassing time-domain, frequency-domain, and statistical characteristics derived from the detected landmarks.

The decision-making process of ML models is often described as a “black box” [38]. We therefore used the SHAP method to interpret the LightGBM model predictions. From the SHAP analysis, we found that the frequency domain feature: the standard deviation of the dominant frequency of the PPG cardiac cycle, followed by the standard deviation of the systolic rise phase width at 33% of the PPG cardiac cycle, as well as the PPG cardiac cycle second-derivative amplitude ratio feature and the PPG cardiac cycle diastolic phase amplitude ratio feature derived from the dicrotic notch and diastolic phase endpoint (see Figure 4 and Table 1), were the most influential features contributing to the model’s predictions.

Shelley et al. [4] demonstrated that venous pulsations of central venous origin can cause high variability in the diastolic phase of the PPG waveform. Moreover, the venous waveform can significantly enlarge the diastolic portion of the PPG signal, sometimes reaching up to 40% of the pulse’s total amplitude [7]. Consistent with these findings, our study found that five of the top 20 features contributing most to model prediction were derived from the diastolic phase of the PPG waveform (see Fig. 4 and Table 1).

This study has several limitations. First, we trained and tested our ML model on a single-center dataset (MLORD); an external validation using a large dataset is needed to assess the generalizability of the model. Second, our analysis was limited to PPG features and incorporating clinically useful features from additional non-invasive human biosignals, such as electrocardiogram (ECG) waveforms, may further improve model performance. Third, in this study, we did not investigate the potential of PPG features to identify patients with elevated CVP based on their recording location (finger, wrist, forearm, or ear); future studies should explore whether incorporating site-specific PPG measurements can improve model performance and generalizability.

## Conclusions

In this study, we developed a ML model (LightGBM) using PPG waveform features to identify patients with elevated CVP. The model’s performance was evaluated using a large perioperative dataset (MLORD). The LightGBM model effectively identified patients with elevated CVP and demonstrated strong, well-balanced performance, achieving an area under the receiver operating characteristic curve (AUC) of 0.79 and an accuracy of 0.71. Although these preliminary results are promising and establish a foundation for noninvasive CVP assessment, further validation using larger and more diverse multicenter datasets is needed to confirm the model’s generalizability and real-world applicability.

## Data Availability

The interested parties may reach out to the first author at or the corresponding author at to request access to the MLORD dataset and the PPG waveform featurization tool.

## Acknowledgements

This work was supported by the National Institutes of Health (NIH): R01EB029751 and R01HL144692.

## Data Accessibility

The interested parties may reach out to the first author at or the to request access to the MLORD dataset and the PPG waveform featurization tool.

## Author contributions statement

R.P. contributed to study design, data analysis, conceptualization, and manuscript preparation. A.R. and J.N.C. contributed to study design and review & editing of the manuscript. A.B. contributed to study design, supervision of the study, review & editing of the manuscript, and conceptualization. M.C. contributed to study design, supervision of the study, review & editing of the manuscript, funding acquisition, and conceptualization. All authors have read and approved the final version of the manuscript.

## Declaration of competing interest

Dr. Cannesson is a consultant for Edwards Lifesciences and Masimo Corp, and has funded research from Edwards Lifesciences and Masimo Corp. He is also the founder of Sironis and Perceptive Medical and he owns patents and receives royalties for closed loop hemodynamic management technologies that have been licensed to Edwards Lifesciences. All the remaining authors declare no conflict of interest.

## Notes

### Author Declarations

Institutional Review Board of University of California, Los Angeles gave ethical approval for this work.

